# Prognostic frailty-based determinants of long-term mortality in older patients with newly diagnosed multiple myeloma

**DOI:** 10.1101/2024.08.15.24312039

**Authors:** Mariya Muzyka, Silvia Ottaviani, Irene Caffa, Tommaso Bonfiglio, Erica Parisi, Ana Guijarro, Luca Tagliafico, Roberto Lemoli, Marta Ponzano, Cristina Marelli, Alessio Signori, Alessio Nencioni, Michele Cea, Fiammetta Monacelli

**Author notes:** To whom correspondence should be addressed: Dr. Alessio Nencioni,; Dr. Michele Cea,; Dr. Fiammetta Monacelli. Share equal senior authorship (Alessio Nencioni, Michele Cea, Fiammetta Monacelli).

## Abstract

**Introduction:** Multiple myeloma (MM) is a plasma cell neoplasm predominantly diagnosed in older adults. However, the significance of defining patient frailty, as well as identifying the most suitable and reliable tools for its assessment, remains to be firmly established.

**Materials and Methods:** This retrospective observational study investigated 36 patients, aged 65 or older, who underwent Comprehensive Geriatric Assessment (CGA). The average patient age was 76 (SD 6.22), with 33.3% being female. Patients were evaluated using the International Myeloma Working Group Frailty Index (IMWG-FI) and the 40-item Rockwood’s Frailty Index (FI) at the Oncogeriatrics clinic of the IRCCS Polyclinic San Martino Hospital, Genoa, Italy, between December 2017 and August 2021. Laboratory, cancer-specific, demographic, and clinical variables were collected. Survival analysis and frailty comparison were conducted using Stata version 17.0.

**Results:** Stepwise multivariate analysis identified Numerical Rating Scale (NRS) (HR 1.40, 95% CI 1.09-1.78, p=0.008) and Rockwood’s Frailty Index (FI) (HR 2.23, 95% CI 1.29-3.87, p=0.004) as significant prognostic predictors, adjusted for sex, ISS stage, and multimorbility. Comparison between Rockwood’s FI and IMWG-FI using Spearman correlation coefficient showed no statistically significant correlation (r=0.268, p=0.114). Multivariate Cox model, adjusting for sex, International Staging System (ISS) stage, and Cumulative Illness Rating Scale (CIRS) comorbidity index, demonstrated the superior predictive ability of Rockwood’s FI over IMWG-FI (C-index 0.775 vs 0.749).

**Discussion:** the 40-item Rockwood FI emerges as a valuable tool for prognostication in old MM patients, surpassing the traditional IMWG-FI in predictive accuracy, emphasizing the importance of a comprehensive approach considering both disease-specific and patient-related factors.

## Introduction

Multiple myeloma (MM) is a plasma cells tumor that typically occurs in older patients: the median age at the time of diagnosis is 69 years, and more than 30% of these are older than 75 years [1]. As a result, given the global increase in the elderly population, the incidence of MM has constantly increased in recent years [2], [3]. Importantly, old age patients represent a highly heterogeneous population with different proportions of multimorbid and frail adults, who are more vulnerable to adverse clinical outcomes; such features often raise uncertainties about the clinical benefit derived from cancer treatments in these patients [4]. In such a context, although clinical benefits in terms of increased Progression Free Survival (PFS) and Overall Survival (OS) derived from novel therapies (such as immunomodulators, proteasome inhibitors, and monoclonal antibodies) and supportive care availability, age still remains the major determinant of outcomes, which obviously gets worst decade by decade [5], [6], [7], [8].

As a result, great caution should be used in the management of elderly patients with several clinical scores developed for the identification of frail and unfit subjects. However, chronological age and performance status alone are insufficient to classify patients’ frailty [9], [10], [11] which identifies a geriatric syndrome characterized by impaired biological and functional reserves that reduce the ability to respond to stressors [12]. To date, although two key relevant frailty constructs have been developed, such as Linda Fried’s physical frailty phenotype [13] and Kenneth Rockwood’s deficit accumulation model [14] both of which address the clinical complexity of older adults, frailty assessment in clinical practice still remains challenging due to heterogeneity and complexity of its measurement. At diagnosis, at least 1/3 of MM patients have some degree of frailty [15] and growing evidence suggests that it is associated with reduced therapeutic response, increased toxicity, worse survival [16], and a higher risk of treatment discontinuation [17].

The International Myeloma Working Group Frailty Index (IMWG-FI) represents the currently most used system to evaluate MM patients’ fitness who are defined as “fit”, “intermediate fit” or “fragile”; a significant impact of this evaluation on both PFS and OS has been demonstrated [18]. However, Scheepers et al. have recently shown the accuracy and appropriateness of a comprehensive geriatric assessment (CGA) for the identification of frailty in older adults with hematologic malignancies: 68% (range 25-76%) of patients according to CGA assessment resulted in being frail, compared with 45% defined by other approaches [16]. Since, older patients with geriatric disabilities have significant clinical benefits, in terms of enhanced responses and reduced adverse events occurrence, from a timely frailty status assessment, the CGA represents the most accurate system to define fitness and thus for selecting optimal strategies for elderly patients [19], [20].

In line with these data, Rockwood’s Frailty Index (FI) has been applied to patients with myelodysplastic syndromes demonstrating that age-adjusted IPSS-R score, FI and Charlson Comorbidity Index were independent prognostic determinants for overall survival. Indeed, frailty and comorbidity scores resulted in improved IPSS-R risk stratification by 30% and 5%, respectively [21]. Conversely, Mian et al., applied the frailty index paradigm to a cohort of MM patients and observed that each 10% increase in frailty index was associated with a 16% increase in the risk of death. The estimated median overall survival of frail patients was 26.8 months, compared with 43.7 months for those who were not frail [22]. Similarly, in a cohort of 3807 U.S. veterans, aged 65 or older, Patel et al. observed that a higher FI score was associated with higher mortality with hazard ratios of 1.33, 1.97, 2.86 and 3.22 for prefrail, mildly frail, moderately frail, and severely frail individuals, respectively [23].

Given this background, we retrospectively compared the prognostic accuracy of IMWG-FI and a 40-item Rockwood FI [24] to predict overall survival (OS) in a cohort of newly diagnosed non-transplant eligible MM patients.

## Materials and Methods

### Patient population and Study design

This retrospective observational study included 54 patients with non-transplant eligible MM, referred to the Oncogeriatrics clinic of the IRCCS Polyclinic San Martino Hospital, Genoa, Italy between December 2017 and August 2021. Inclusion criteria were patients aged 65 or older newly diagnosed with MM not eligible for high-dose therapies, who underwent a CGA at diagnosis and received treatment at the Oncogeriatrics clinic during the specific period, with complete clinical data available. Exclusion criteria included age below 65 years, patients with new diagnosis of MM transplant eligible, monoclonal gammopathy of undetermined significance (MGUS) or smoldering myeloma, absence of CGA or complete clinical data, and treatment received outside the specific clinic. The study wa apporved by the IRB CERA University of Genoa ,N.2024,54, Italy. Authors did not access to information that could identify individual participants during or after data collection

### Laboratory and cancer-specific variables

Disease-related variables included SLiM-CRAB criteria present at diagnosis [20], histotype, cytogenetic risk status assessed by fluorescence in situ hybridization (FISH), bone disease, extramedullary disease, flow cytometric data from bone marrow aspirates (BM), disease staging system according to Durie-Salmon (D-S) [25] and to the International Staging System (ISS) [26], if available. Laboratory variables included blood count, creatinine, urea, coagulation profile (international normalized ratio, prothrombin time, partial thromboplastin time), calcium, serum albumin, total serum proteins, total and direct bilirubin, alanine transaminase, aspartate transaminase, beta2-microglobulin, lactate dehydrogenase, serum free light chain ratio, M component, 24-hour proteinuria, and 24-hour Bence Jones proteinuria.

### Demographic and Clinical Assessments

Demographic data included age and sex. Following an initial frailty screening using the G8 tool [27] (with, a cut-off of ≤14), each patient underwent a CGA at MM diagnosis, including cognitive status using Mini-Mental State Examination (MMSE) and Clock Drawing test according to Shulman (CDT) [28], [29]. Nutritional risk was assessed using Mini Nutritional Assessment (MNA) [30]; functional autonomy with Barthel Index and Instrumental Activity of Daily Living (IADL) [31], [32]: multimorbidity with Cumulative Illness Rating Scale (CIRS) [33]; psycho-affective status with 15-item Geriatric Depression Scale (GDS) [34]; gait and risk of falling with the Tinetti scale [35] and pain evaluation with the Numerical Rating Scale (NRS) [36]. Social vulnerability was assessed with the Gijon scale [37] and sarcopenia and physical performance with Hand Grip (HG) test (using a GIMA 28791 Smedley dynamometer), SARC-F scale [38] and the Timed-Up and Go test (TUG) [39], [40]. Polypharmacy was assessed by counting the number of drugs [41] and quality of life was assessed with the EuroQol instrument (EQ-5D) [42]. A CGA cut-off>3 defined frailty [43].

For frailty stratification, the IMWG-FI and the Rockwood’s FI were calculated. IMWG score includes the patient’s age and CGA measurements including Activities of Daily Living (ADL), Instrumental Activities of Daily Living (IADL), and comorbidity (Charlson Comorbidity Index (CCI) [44]). The IMWG scoring system indicates frailty levels: fit (score = 0), intermediate-fit (score = 1), and frail (score ≥2).

Rockwood’s FI considers 40 health deficits, with cumulative scores indicating frailty levels: fit (score≤0.08); prefrail (score: 0.09<pre frail <0.24); frail (score ≥0.25). See **S1** for tool details. Time to death from all causes between the first geriatric visit at MM diagnosis and death was recorded. The date of censorship was 08/31/2022. Data were accessed for research purposes between January 2021 and agust 2022.

### Statistical analysis

Results were described using mean and standard deviation (SD) or median and interquartile range (IQR) or absolute frequency (N) and relative frequency (%), according to the distribution. The normal distribution hypothesis was checked through the Shapiro-Wilk test. The Spearman correlation coefficient assessed the correlation between IMWG-FI and Rockwood’s FI. Characteristics of alive or deceased patients were compared through the t-test, the Wilcoxon sum-of-ranks test, or the chi-squared test, according to the distribution. Kaplan-Meier survival curve was used to assess time to death and univariate Cox proportional hazards models were performed to evaluate the impact of the variables on mortality. A multivariate Cox model adjusted for sex, stage ISS, and CIRS comorbidity index with backward stepwise selection was run including variables with a p-value<0.05 in the univariate analysis, excluding variables with higher degree of collinearity [45].

A multivariate model including all the selected variables with backward stepwise selection and adjusting for age was performed for sensitivity analysis. The final stepwise multivariate model was adjusted for age. The proportional hazard assumption of the final multivariate model was checked using Schoenfeld residuals. Rockwood’s FI and IMWG-FI were compared using the C-index, the AIC and BIC values, adjusting for sex, stage ISS, and CIRS comorbidity index and the same analysis was performed to assess sensitivity.

All tests were two-sided and p-values <0.05 were considered statistically significant. All analyses used Stata version 17.0 (Stata Corporation).

## Results

### Cohort Characteristics

Between December 2017 and August 2021, 54 consecutive newly diagnosed MM patients who were non-transplant eligible, were analyzed. Of 54 admitted patients, 36 were included in the final analysis. 4 patients were excluded for a diagnosis of MGUS, and 1 patient was excluded for a diagnosis of smoldering MM; a further 13 patients were excluded because the CGA assessment was performed far after the diagnosis of MM. Baseline characteristics detailed in Table 1, showed a mean age of 76 (±6.22) years (range: 65-92 years) with 33% aged over 80 and 67% male. Most patients had IgG histotype at diagnosis (64%); the most represented disease stages were IIA and IIIA according to Durie-Salmon and II and III based on ISS system, a minor prevalence was observed for stage I. A standard cytogenetic risk was observed in the majority of included patients (80%) with at least one bone lesion observed at the time of diagnosis in almost all cases. Extramedullary disease was observed in only 2 patients. Detailed blood and urine tests and cytofluorimetric data are illustrated in S2.

**Table 1.** Baseline characteristics of the population.

|  | **Overall,**<br>**N=36 (100%)** | **Dead,**<br>**N=14 (39%)** | **Alive,**<br>**N=22 (61%)** | **p-value** |
| --- | --- | --- | --- | --- |
| Female sex | 12 (33.33%) | 3 (21.43%) | 9 (40.91%) | 0.227 |
| Age | 76 (6.22) | 78.21 (6.76) | 74.59 (5.56) | 0.089 |
| IMWG-FI, median [IQR] | 1 [0-2] | 1.5 [1-3] | 1 [0-2] | 0.367 |
| G8 ($\leq 14$), median [IQR] | N=35<br>13 [12-15] | N=13<br>12 [11-14] | N=22<br>13.25 [12-16] | 0.130 |
| MMSE, median [IQR] | 28 [26.6-29] | 27.85 [26.4-28.7] | 28 [27-29] | 0.442 |
| Clock drawing test, median [IQR] | N=35<br>2 [1-2] | N=14<br>2 [1-3] | N=21<br>1 [1-2] | 0.137 |
| MNA | 23.93 (3.68) | 22.54 (3.37) | 24.82 (3.66) | 0.069 |
| IADL, median [IQR] | 7 [7-8] | 8 [7-8] | 8 [8-8] | **0.017** |
| Barthel, median [IQR] | 100 [95-100] | 97.5 [90-100] | 100 [95-100] | 0.381 |
| CIRS comorbidity index | 4.29 (1.92) | 4.74 (2.11) | 4 (1.77) | 0.263 |
| GDS, median [IQR] | N=35<br>3 [1-4] | N=14<br>3.5 [1-6] | N=21<br>3 [2-3] | 0.388 |
| Tinetti test, median [IQR] | N=34<br>26 [22-28] | N=13<br>23 [21-27] | N=21<br>26 [2-28] | 0.276 |
| NRS, median [IQR] | 4 [0-5] | 5 [3-6] | 2 [0-5] | 0.057 |
| Gjion | 8.75 (2.66) | 8.57 (2.38) | 8.86 (2.87) | 0.753 |
| TUG, median [IQR] | N=34<br>9.5 [7.2-12] | N=13<br>10 [7.2-12] | N=21<br>8.5 [8-11] | 0.644 |
| HG | 27.92 (9.16) | 29.76 (8.32) | 26.75 (9.67) | 0.345 |
| SARC-F | N=20<br>2.85 (2.39) | N=8<br>3.13 (2.42) | N=12<br>2.67 (2.46) | 0.686 |
| cut off CGA ($\geq 3$) | 3.19 (2.15) | 3.71 (2.16) | 2.86 (2.12) | 0.253 |
| Polypharmacy | 6.22 (3.45) | 6.93 (3.79) | 5.77 (3.22) | 0.334 |
| Rockwood's FI, median [IQR] | 0.18 [0.11-0.23] | 0.21 [0.18-0.27] | 0.16 [0.1-0.2] | 0.066 |
| EuroQoL | N=35<br>0.70 (0.17) | N=13<br>0.63 (0.20) | N=22<br>0.74 (0.14) | 0.072 |
| RT, yes | N=32<br>10 (31.25%) | N=14<br>4 (28.57%) | N=18<br>6 (33.33%) | 0.773 |
| Stage D-S | N=35 | N=14 | N=21 | **0.048** |
| IA | 5 (14.29%) | 0 (0.00%) | 5 (23.81%) |  |
| IIA | 11 (31.43%) | 6 (42.86%) | 5 (23.81%) |  |
| IIIA | 12 (34.29%) | 7 (50.00%) | 5 (23.81%) |  |
| IIIB | 7 (20.00%) | 1 (7.14%) | 6 (28.57%) |  |
| Stage ISS |  |  |  | 0.221 |
| I | 8 (22.22%) | 1 (7.14%) | 7 (31.82%) |  |
| II | 13 (36.11%) | 6 (42.86%) | 7 (31.82%) |  |
| III | 15 (41.67%) | 7 (50.00%) | 8 (36.36%) |  |

(i) the entire population; (ii) dead patients; (iii) alive patients. Results are presented through mean and standard deviation (SD) and absolute frequency (N) and relative frequency (%), for continuous and categorical variables respectively, unless otherwise specified. P-values of the t-test, the Wilcoxon sum-of-ranks test, or the chi-squared test are presented to compare (ii) and (iii), according to the distribution.
Abbreviation list. MMSE: Mini-Mental State Examination; MNA: Mini Nutritional Assessment; IADL: Instrumental Activity of Daily Living; CIRS: Cumulative Illness Rating Scale; GDS: 15-item Geriatric Depression Scale; NRS: Numerical Rating Scale; HG: Hand Grip; TUG: Timed Up and Go test; Rockwood's FI: Rockwood's Frailty Index; RT: radiotherapy; D-S: Durie-Salmon; ISS: International Staging System. HR: hazard ratio; CI: confidence interval; IQR: interquartile range.

### CGA and Frailty assessment at baseline

On the basis of CGA, almost half of the patients (44%) were at higher malnutrition risk, 39% had impaired instrumental activities and had a mean of 4 comorbidities and 5 drugs respectively. We subsequently compared the fitness of included patient by measuring their frailty scores derived from the IMWG-FI and Rockwood’s FI. As shown in Table 3, the IMWG-FI mean score was 1 IQR 0-2), and the Rockwood FI mean score was 0.18 (IQR 0.11-0.23). These data suggest an imbalance between these systems: indeed, 64% of patients resulted as pre-frail based on Rockwood’s FI, while most patients were classified as frail according to IMWG-FI.

### Outcomes

According to the censorship data, 14 patients (39%) died, with 14% passing away within the first year. Specifically, 3 patients died of disease progression and related complications, 2 of COVID-19 pneumonia, 1 of acute pulmonary edema, and the cause of death for the remaining 8 patients was unknown. Fig 1 illustrates the Kaplan-Meier curve, depicting the mortality trend within the cohort over 5.77 years. As detailed in Table 1, surviving patients demonstrated higher levels of functional autonomy in instrumental activities (p=0.017), and lower disease severity according to the Durie-Salmon score (p=0.048).

Following a preliminary univariate survival analysis, the stepwise multivariate analysis identified NRS (HR 1.40, 95% CI 1.09-1.78, p=0.008) and Rockwood’s FI (HR 2.23, 95% CI 1.29-3.87, p=0.004) as significant predictors, along with age (HR 0.15, 95% CI 0.03-0.75, p=0.021) and advanced stage of disease (HR 6.09, 95% CI 1.36-27.21, p=0.018). Sex, ISS stage, and CIRS comorbidity index were included in the model as adjustments (Table 2).

To compare the predictive ability of the model between Rockwood’s FI and IMWG-FI, we first evaluated the correlation between IMWG-FI and Rockwood’s FI using the Spearman correlation coefficient, which was found not to be statistically significant: r=0.268 (p=0.114). Subsequently, a multivariate Cox model was built with the variable of interest (IMWG-FI vs Rockwood’s FI) and adjustment for sex, ISS stage, and CIRS comorbidity index. AIC, BIC, and C-index were then calculated for comparison. The data presented in Table 3 indicate that the model including Rockwood’s FI showed superior predictive ability regarding the survival of older patients with MM, with equal stage of disease and multimorbidity burden (C-index 0.775 vs 0.749). Sensitivity analysis, adjusted for age, yielded consistent results with those reported above.

**Fig 1.**
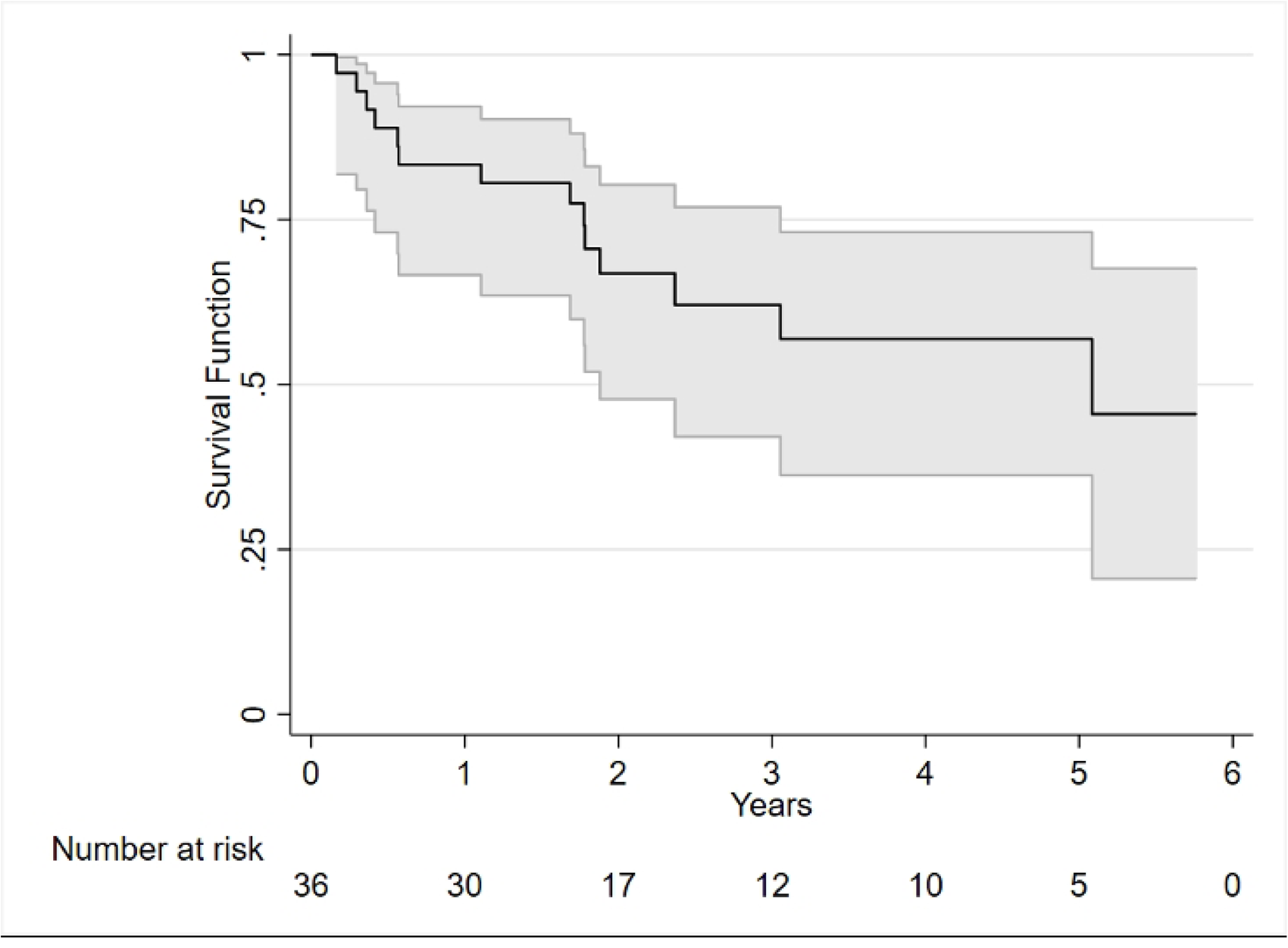
Kaplan-Meier graph estimating the survival probability over time.

**Table 2.** Univariate and backward stepwise multivariate Cox proportional hazards model for death.

|  | Univariate |  | Multivariate stepwise,<br>including sex, stage ISS,<br>and CIRS t0<br>(comorbidity) |  |
| --- | --- | --- | --- | --- |
|  | HR(95% CI) | p-value | HR(95% CI) | p-value |
| Sex (female vs. male) | 0.55 (0.15-1.96) | 0.354 | 0.15 (0.03-0.75) | **0.021** |
| Age | 1.10 (1.01-1.19) | **0.022\*** | - | - |
| IMWG-FI | 1.67 (1.09-2.57) | **0.019** | - | - |
| G8 ($\leq 14$) (N=35) | 0.76 (0.60-0.95) | **0.016** | - | - |
| MMSE | 0.90 (0.70-1.14) | 0.368 | - | - |
| Clock drawing test (N=35) | 1.43 (0.90-2.27) | 0.127 | - | - |
| MNA | 0.81 (0.69-0.95) | **0.008\*** | - | - |
| IADL | 0.73 (0.56-0.96) | **0.024\*** | - | - |
| Barthel index | 0.96 (0.91-1.01) | 0.158 | - | - |
| CIRS comorbidity index | 1.21 (0.94-1.54) | 0.137 | - | - |
| GDS (N=35) | 1.27 (1.04-1.55) | **0.021** | - | - |
| Tinetti (N=34) | 0.92 (0.81-1.04) | 0.195 | - | - |
| NRS | 1.25 (1.02-1.54) | **0.034** | 1.40 (1.09-1.78) | **0.008** |
| Gjion | 1.05 (0.86-1.29) | 0.626 | - | - |
| TUG (N=34) | 1.07 (0.94-1.22) | 0.311 | - | - |
| HG | 1.01 (0.96-1.06) | 0.781 | - | - |
| SARC F (N=20) | 1.10 (0.80-1.51) | 0.556 | - | - |
| cut off CGA ($\geq 3$) | 1.36 (1.03-1.79) | **0.027\*** | - | - |
| Polypharmacy | 1.12 (0.96-1.31) | 0.139 | - | - |
| Rockwood's FI (0.1 increase) | 1.57 (1.07-2.31) | **0.021** | 2.23 (1.29-3.87) | **0.004** |
| EuroQoL (0.1 increase) (N=35) | 0.75 (0.55-1.02) | 0.066 | - | - |
| RT (yes vs. no) (N=32) | 0.68 (0.21-2.22) | 0.525 | - | - |
| Stage D-S (IA or IIA vs. IIIA or IIIB) (N=35) | 0.59 (0.20-1.76) | 0.348 | - | - |
| Stage ISS (I or II vs. III) | 0.36 (0.12-1.10) | 0.072 | 6.09 (1.36-27.21) | **0.018** |

HR: hazard ratio; CI: confidence interval. \* Excluded from the multivariate model due to the high correlation with another variable included.

**Table 3.** AIC, BIC, and C-index for the comparison between Rockwood’s FI and IMWG-FI.

|  | AIC | BIC | C-index |
| --- | --- | --- | --- |
| Model A | 83.45 | 89.78 | 0.7745 |
| Model B | 88.35 | 94.68 | 0.7493 |

Model A: multivariate Cox model for survival time, including Rockwood's FI and adjusting for sex, stage ISS, and CIRS comorbidity index; model B: multivariate Cox model for survival time, including IMWG-FI and adjusting for sex, stage ISS, and CIRS comorbidity index.

## Discussion

MM is predominantly a disease of old age adults, thus geriatric assessment is emerging as crucial to identify vulnerabilities and frailty, that compromise treatment outcomes and susceptibility to adverse events derived from different therapies. Older adults with MM represent a growing demographic, warranting personalized treatments based on biological health status to improve survival, maintenance of functional reserve, and quality of life. While existing evidence has focused on cancer-related prognostic variables and validated myeloma-specific scores to delineate fitness versus frailty status in older adults, the full understanding of the interplay between frailty and cancer survival remains incomplete, as does the systematic application of such scores in routine practice. In assessing older adults with MM, both cancer-specific and patient-specific factors may undermine functional reserve, influencing prognosis, complicating treatment decisions, toxicity assessment, and shared decision-making processes.

This study paves the way to integrate laboratory and MM-specific variables with patients’ frailty, employing a methodologically robust stratification to predict OS in a real-world cohort of older newly diagnosed MM patients non eligible for HD-therapies. Our findings demonstrated that the 40-item Rockwood FI was significantly associated with OS, outperforming the gold standard IMWG-FI along with a comprehensive set of MM-specific variables. These original findings underscore how patient-based frailty stratification impacts MM clinical management, progression, and outcomes. Notably, the study population was aged over 75 years, providing valuable insights into frailty measurement applicability in very old individuals.

Currently available clinical scores, including IMWG-FI, classify patients as ‘fit’, ‘intermediate-fit’ or ‘frail’, based on chronological age, with patients over 80 automatically classified as frail [46]. However, with the increasing clinical heterogeneity of aging populations, relying solely on chronological age risks undertreating patients who may benefit from more intensive therapeutic regimens. In such a context, our frailty stratification identified vulnerable patients as at intermediate risk, including one-third of the oldest old and, notably, our study revealed that chronological age alone did not correlate with an increased likelihood of frailty.

Moreover, by incorporating a series of disease-related prognostic factors into the predictive model, our approach captures the clinical complexity of MM and strengthens the prognostic ability of Rockwood’s FI which, in our hands, outperformed both MM-related factors and IMWG-FI.

Similarly, Stege et al. demonstrated that older patients aged 75 years and older with newly diagnosed MM categorized as frail based on IMWG-FI scoring had comparable outcomes to patients classified as intermediate fit [43]. Moreover, Murillo et al. underscored that frailty-based stratification with Fried’s physical phenotype outperformed IMWG-FI classification, suggesting that biological age rather than chronological age plays a critical role in informing outcomes [47].

Mian et al. utilized Rockwood’s deficit accumulation model on a large sample of older patients without cancer to derive a frailty index applicable to MM patients. This final 25-item model was associated with OS even after adjusting for chronological age, emphasizing the differences between biological and chronological age [20]. Particularly, in the cohort of newly diagnosed MM, the mean frailty index was 0.28, and each 10% increase in frailty index was associated with a 16% increased risk of death, with a median OS estimated at 26.8 months. In our study, a 0.1 increase in Rockwood’s FI resulted in a 112% increase in the risk of death at the end of the observation period.

Our findings contrast with those reported by Tyczynska et al., who concluded that the management of patients over 75 was not dependent on frailty assessment [48]. However, in that cohort, most patients had a relatively good performance status, potentially undermining the generalizability of the findings due to the lack of robust frailty stratification.

It could be hypothesized that the cumulative burden of clinical deficits, including comorbidity, functional impairment, mental status, general health status, and quality of life, may be related to the overall burden of MM, ultimately increasing the likelihood of mortality. Rockwood’s FI captures the dynamic changes of frailty when superimposed with major stressors such as MM and cancer therapies, influencing functional reserve, remission, disease-free progression, and outcomes such as long-term mortality, disability, and quality of life.

Our study has important limitations, including single-center enrollment and its retrospective nature. The study sample is skewed towards males although consistent with literature data [49].

Additionally, the small number of patients and “death” events limited our ability to establish the prognostic value of the 40-item Rockwood FI. Furthermore, other important outcomes such as PFS, therapeutic regimen, or hospitalization were not reported, warranting further in-depth analysis.

Despite these limitations, the study offers a comprehensive evaluation of old-age MM patients, adopting a methodologically robust frailty assessment. Focusing on older adults aged over 75 years, this study addresses the unique needs and challenges faced by this growing demographic within the MM patient population. Our findings, though preliminary, have real-world applicability, providing actionable insights that can inform and enhance clinical practice.

In such a complex therapeutic landscape, with the availability of increasingly effective treatments, such as bispecific antibodies and CAR-T strategies, the in-depth analysis of MM patient’s frailty is becoming imperative.

In conclusion, the 40-item Rockwood FI frailty stratification model, by demonstrating effectiveness in predicting time to death in older patients with MM, establishes itself as a valuable and innovative prognostic tool. Our study underscores the importance of precise patient selection, highlighting the limitations of relying solely on chronological age for patient phenotyping. Rockwood’s FI can capture the dynamic nature of frailty, heavily influenced by the disease burden of MM. This highlights its significance in refining prognostic assessments and guiding personalized treatment strategies for older adults affected by this hematologic malignancy.

## Data Availability

ll relevant data are within the manuscript and its Supporting Information files.

## Acknowledgments

We would like to thank all the authors for their contributions to this research. We are also grateful for the support provided by the University of Genoa and IRCCS Polyclinic San Martino Hospital.

## References

[1] S. V. Rajkumar, «Multiple myeloma: 2022 update on diagnosis, risk stratification, and management», Am. J. Hematol., vol. 97, fasc. 8, pp. 1086–1107, ago. 2022, doi: 10.1002/ajh.26590.

[2] A. J. Cowan et al., «Global Burden of Multiple Myeloma: A Systematic Analysis for the Global Burden of Disease Study 2016», JAMA Oncol., vol. 4, fasc. 9, p. 1221, set. 2018, doi: 10.1001/jamaoncol.2018.2128.

[3] P. S. Rosenberg, K. A. Barker, eW. F. Anderson, «Future distribution of multiple myeloma in the United States by sex, age, and race/ethnicity», Blood, vol. 125, fasc. 2, pp. 410–412, gen. 2015, doi: 10.1182/blood-2014-10-609461.

[4] A. Palumbo et al., «Personalized therapy in multiple myeloma according to patient age and vulnerability: a report of the European Myeloma Network (EMN)», Blood, vol. 118, fasc. 17, pp. 4519–4529, ott. 2011, doi: 10.1182/blood-2011-06-358812.

[5] S. K. Kumar et al., «Improved survival in multiple myeloma and the impact of novel therapies», Blood, vol. 111, fasc. 5, pp. 2516–2520, mar. 2008, doi: 10.1182/blood-2007-10-116129.

[6] M.-V. Mateos et al., «Bortezomib, melphalan, and prednisone versus bortezomib, thalidomide, and prednisone as induction therapy followed by maintenance treatment with bortezomib and thalidomide versus bortezomib and prednisone in elderly patients with untreated multiple myeloma: a randomised trial», Lancet Oncol., vol. 11, fasc. 10, pp. 934–941, ott. 2010, doi: 10.1016/S1470-2045(10)70187-X.

[7] S. Pozzi et al., «Survival of multiple myeloma patients in the era of novel therapies confirms the improvement in patients younger than 75 years: a population-based analysis», Br. J. Haematol., vol. 163, fasc. 1, pp. 40–46, ott. 2013, doi: 10.1111/bjh.12465.

[8] C. Pawlyn et al., «The relative importance of factors predicting outcome for myeloma patients at different ages: results from 3894 patients in the Myeloma XI trial», Leukemia, vol. 34, fasc. 2, pp. 604–612, feb. 2020, doi: 10.1038/s41375-019-0595-5.

[9] A. Palumbo et al., «Geriatric assessment predicts survival and toxicities in elderly myeloma patients: an International Myeloma Working Group report», Blood, vol. 125, fasc. 13, pp. 2068– 2074, mar. 2015, doi: 10.1182/blood-2014-12-615187.

[10] A. Larocca et al., «Patient-centered practice in elderly myeloma patients: an overview and consensus from the European Myeloma Network (EMN)», Leukemia, vol. 32, fasc. 8, pp. 1697–1712, ago. 2018, doi: 10.1038/s41375-018-0142-9.

[11] E. Soto-Perez-de-Celis, D. Li, Y. Yuan, Y. M. Lau, e A. Hurria, «Functional versus chronological age: geriatric assessments to guide decision making in older patients with cancer», Lancet Oncol., vol. 19, fasc. 6, pp. e305–e316, giu. 2018, doi: 10.1016/S1470-2045(18)30348-6.

[12] W. De Alfieri, S. Costanzo, T. Borgogni, «Biological resilience of older adults versus frailty», Med. Hypotheses, vol. 76, fasc. 2, pp. 304–305, feb. 2011, doi: 10.1016/j.mehy.2010.11.028.

[13] L. P. Fried et al., «Frailty in Older Adults: Evidence for a Phenotype», J. Gerontol. A. Biol. Sci. Med. Sci., vol. 56, fasc. 3, pp. M146–M157, mar. 2001, doi: 10.1093/gerona/56.3.M146.

[14] A. B. Mitnitski, A. J. Mogilner, eK. Rockwood, «Accumulation of Deficits as a Proxy Measure of Aging», Sci. World J., vol. 1, pp. 323–336, 2001, doi: 10.1100/tsw.2001.58.

[15] M. Engelhardt et al., «A concise revised Myeloma Comorbidity Index as a valid prognostic instrument in a large cohort of 801 multiple myeloma patients», Haematologica, vol. 102, fasc. 5, pp. 910–921, mag. 2017, doi: 10.3324/haematol.2016.162693.

[16] E. R. M. Scheepers, A. M. Vondeling, N. Thielen, R. Van Der Griend, R. Stauder, e M. E. Hamaker, «Geriatric assessment in older patients with a hematologic malignancy: a systematic review», Haematologica, vol. 105, fasc. 6, pp. 1484–1493, giu. 2020, doi: 10.3324/haematol.2019.245803.

[17] A. Laroccae A. Palumbo, «How I treat fragile myeloma patients», Blood, vol. 126, fasc. 19, pp. 2179–2185, nov. 2015, doi: 10.1182/blood-2015-05-612960.

[18] A. Palumbo et al., «Revised International Staging System for Multiple Myeloma: A Report From International Myeloma Working Group», J. Clin. Oncol. Off. J. Am. Soc. Clin. Oncol., vol. 33, fasc. 26, pp. 2863–2869, set. 2015, doi: 10.1200/JCO.2015.61.2267.

[19] N. Kint e M. Delforge, «Concise review - Treatment of multiple myeloma in the very elderly: How do novel agents fit in?», J. Geriatr. Oncol., vol. 7, fasc. 5, pp. 383–389, set. 2016, doi: 10.1016/j.jgo.2016.08.001.

[20] S. Zweegman, M. Engelhardt, A. Larocca, e EHA SWG on ‘Aging and Hematology’, «Elderly patients with multiple myeloma: towards a frailty approach?», Curr. Opin. Oncol., vol. 29, fasc. 5, pp. 315–321, set. 2017, doi: 10.1097/CCO.0000000000000395.

[21] R. Buckstein et al., «Patient-related factors independently impact overall survival in patients with myelodysplastic syndromes: an MDS-CAN prospective study», Br. J. Haematol., vol. 174, fasc. 1, pp. 88–101, lug. 2016, doi: 10.1111/bjh.14033.

[22] H. S. Mian, T. M. Wildes, e M. A. Fiala, «Development of a Medicare Health Outcomes Survey Deficit-Accumulation Frailty Index and Its Application to Older Patients With Newly Diagnosed Multiple Myeloma», *JCO Clin*. Cancer Inform., vol. 2, p. CCI.18.00043, 2018, doi: 10.1200/CCI.18.00043.

[23] B. G. Patel, S. Luo, T. M. Wildes, e K. M. Sanfilippo, «Frailty in Older Adults With Multiple Myeloma: A Study of US Veterans», *JCO Clin*. Cancer Inform., vol. 4, pp. 117–127, feb. 2020, doi: 10.1200/CCI.19.00094.

[24] C. Giannotti et al., «Frailty assessment in elective gastrointestinal oncogeriatric surgery: Predictors of one-year mortality and functional status», J. Geriatr. Oncol., vol. 10, fasc. 5, pp. 716–723, set. 2019, doi: 10.1016/j.jgo.2019.04.017.

[25] B. G. M. Durie S. E. Salmon, «A clinical staging system for multiple myeloma correlation of measured myeloma cell mass with presenting clinical features, response to treatment, and survival», Cancer, vol. 36, fasc. 3, pp. 842–854, set. 1975, doi: 10.1002/1097-0142(197509)36:3<842::AID-CNCR2820360303>3.0.CO;2-U.

[26] P. R. Greipp et al., «International staging system for multiple myeloma», J. Clin. Oncol. Off. J. Am. Soc. Clin. Oncol., vol. 23, fasc. 15, pp. 3412–3420, mag. 2005, doi: 10.1200/JCO.2005.04.242.

[27] S. Rostoft, A. O’Donovan, P. Soubeyran, S. M. H. Alibhai, e M. E. Hamaker, «Geriatric Assessment and Management in Cancer», J. Clin. Oncol., vol. 39, fasc. 19, pp. 2058–2067, lug. 2021, doi: 10.1200/JCO.21.00089.

[28] G. B. Frisoni, R. Rozzini, A. Bianchetti, e M. Trabucchi, «Principal Lifetime Occupation and MMSE Score in Elderly Persons», J. Gerontol., vol. 48, fasc. 6, pp. S310–S314, nov. 1993, doi: 10.1093/geronj/48.6.S310.

[29] K. I. Shulman, D. Pushkar Gold, C. A. Cohen, e C. A. Zucchero, «Clock-drawing and dementia in the community: A longitudinal study», Int. J. Geriatr. Psychiatry, vol. 8, fasc. 6, pp. 487–496, giu. 1993, doi: 10.1002/gps.930080606.

[30] Y. Guigoz, B. Vellas, e P. J. Garry, «Assessing the nutritional status of the elderly: The Mini Nutritional Assessment as part of the geriatric evaluation», Nutr. Rev., vol. 54, fasc. 1 Pt 2, pp. S59–65, gen. 1996, doi: 10.1111/j.1753-4887.1996.tb03793.x.

[31] F. I. Mahoney e D. W. Barthel, «FUNCTIONAL EVALUATION: THE BARTHEL INDEX», Md. State Med. J., vol. 14, pp. 61–65, feb. 1965.

[32] M. P. Lawton e E. M. Brody, «Assessment of older people: self-maintaining and instrumental activities of daily living», The Gerontologist, vol. 9, fasc. 3, pp. 179–186, 1969.

[33] B. S. Linn, M. W. Linn, e L. Gurel, «Cumulative illness rating scale», J. Am. Geriatr. Soc., vol. 16, fasc. 5, pp. 622–626, mag. 1968, doi: 10.1111/j.1532-5415.1968.tb02103.x.

[34] J. A. Yesavage e J. I. Sheikh, «9/Geriatric Depression Scale (GDS): Recent Evidence and Development of a Shorter Version», Clin. Gerontol., vol. 5, fasc. 1–2, pp. 165–173, nov. 1986, doi: 10.1300/J018v05n01_09.

[35] M. E. Tinetti, «Performance-oriented assessment of mobility problems in elderly patients», J. Am. Geriatr. Soc., vol. 34, fasc. 2, pp. 119–126, feb. 1986, doi: 10.1111/j.1532-5415.1986.tb05480.x.

[36] W. W. Downie, P. A. Leatham, V. M. Rhind, V. Wright, J. A. Branco, e J. A. Anderson, «Studies with pain rating scales», Ann. Rheum. Dis., vol. 37, fasc. 4, pp. 378–381, ago. 1978, doi: 10.1136/ard.37.4.378.

37. J. V. García González et al., «[An evaluation of the feasibility and validity of a scale of social assessment of the elderly]», *Aten. Primaria*, vol. 23, fasc. 7, pp. 434–440, apr. 1999.

[38] T. K. Malmstrom, D. K. Miller, E. M. Simonsick, L. Ferrucci, e J. E. Morley, «SARC-F: a symptom score to predict persons with sarcopenia at risk for poor functional outcomes», J. Cachexia Sarcopenia Muscle, vol. 7, fasc. 1, pp. 28–36, mar. 2016, doi: 10.1002/jcsm.12048.

[39] D. Podsiadlo S. Richardson, «The timed “Up & Go”: a test of basic functional mobility for frail elderly persons», J. Am. Geriatr. Soc., vol. 39, fasc. 2, pp. 142–148, feb. 1991, doi: 10.1111/j.1532-5415.1991.tb01616.x.

[40] R. W. Bohannon, «Muscle strength: clinical and prognostic value of hand-grip dynamometry», Curr. Opin. Clin. Nutr. Metab. Care, vol. 18, fasc. 5, pp. 465–470, set. 2015, doi: 10.1097/MCO.0000000000000202.

[41] J. Tjia, S. J. Velten, C. Parsons, S. Valluri, e B. A. Briesacher, «Studies to reduce unnecessary medication use in frail older adults: a systematic review», Drugs Aging, vol. 30, fasc. 5, pp. 285–307, mag. 2013, doi: 10.1007/s40266-013-0064-1.

[42] EuroQol Group, «EuroQol--a new facility for the measurement of health-related quality of life», Health Policy Amst. Neth., vol. 16, fasc. 3, pp. 199–208, dic. 1990, doi: 10.1016/0168-8510(90)90421-9.

[43] C. A. M. Stege et al., «Improving the identification of frail elderly newly diagnosed multiple myeloma patients», Leukemia, vol. 35, fasc. 9, pp. 2715–2719, set. 2021, doi: 10.1038/s41375-021-01162-z.

[44] M. E. Charlson, P. Pompei, K. L. Ales, e C. R. MacKenzie, «A new method of classifying prognostic comorbidity in longitudinal studies: development and validation», J. Chronic Dis., vol. 40, fasc. 5, pp. 373–383, 1987, doi: 10.1016/0021-9681(87)90171-8.

[45] M. M. Mukaka, «Statistics corner: A guide to appropriate use of correlation coefficient in medical research», Malawi Med. J. J. Med. Assoc. Malawi, vol. 24, fasc. 3, pp. 69–71, set. 2012.

[46] M. D’Agostino et al., «Octogenarian newly diagnosed multiple myeloma patients without geriatric impairments: the role of age >80 in the IMWG frailty score», Blood Cancer J., vol. 11, fasc. 4, p. 73, apr. 2021, doi: 10.1038/s41408-021-00464-w.

[47] A. Murillo et al., «Performance of the International Myeloma Working Group myeloma frailty score among patients 75 and older», J. Geriatr. Oncol., vol. 10, fasc. 3, pp. 486–489, mag. 2019, doi: 10.1016/j.jgo.2018.10.010.

[48] A. Tyczyńska et al., «The Real-World Evidence on the Fragility and Its Impact on the Choice of Treatment Regimen in Newly Diagnosed Patients with Multiple Myeloma over 75 Years of Age», Cancers, vol. 15, fasc. 13, p. 3469, lug. 2023, doi: 10.3390/cancers15133469.

[49] S. Bird et al., «Sex Differences in Multiple Myeloma Biology but not Clinical Outcomes: Results from 3894 Patients in the Myeloma XI Trial», Clin. Lymphoma Myeloma Leuk., vol. 21, fasc. 10, pp. 667–675, ott. 2021, doi: 10.1016/j.clml.2021.04.013.

